# Home Blood Pressure and Asymptomatic Intracranial and Extracranial Arterial Stenosis in Hypertension Patients

**DOI:** 10.1101/2024.10.21.24315899

**Authors:** Enheng Cai, Dewei An, Jin Zhang, Xiaofeng Tang, Yan Li, Yan Wang, Dingliang Zhu

## Abstract

**Background:** We aim to analyze the effect of home blood pressure (HBP) level and variability on both asymptomatic intracranial (aICAS) and extracranial arterial stenosis (aECAS).

**Methods:** A total of 534 patients completed HBP measurements (HBP) on at least 3 days. The stenoses of ICAS and ECAS were evaluated by CTA. HBP variability (HBPV) was quantified using the standard deviation and maximum--minimum difference between measures. The association of HBP level and variability with both aICAS and aECAS was evaluated using multivariate logistic regression.

**Results:** Morning HBPV was significantly associated with isolated aICAS, isolated aECAS, and concurrent aICAS and aECAS, respectively, even after being adjusted for HBP level. Besides, HBP were independently related to concurrent aICAS and aECAS (P<0.05). Instead, neither morning nor evening HBP associated with isolated aICAS or aECAS.

**Conclusion:** The findings of this study confirmed the strong correlation of aICAS/aECAS with HBP in the hypertensive population, particularly morning HBPV.

## Introduction

Stroke is one of the common causes of mortality worldwide, causing around 6.17 million deaths every year^1^. Asymptomatic intracranial (aICAS) and extracranial arterial stenosis (aECAS), which are the early phase of symptomatic injury, are considered as the risk factors for stroke and related cardiovascular diseases^2-5^. Notably, the prevalence and risk factors of ICAS and ECAS are different among populations, whereas ICAS is more frequently affected Asians, blacks, and Hispanics, and ECAS is more prevalent among whites.^6^

Elevated blood pressure (BP) is a well-established risk factor for both ischemic and hemorrhagic forms of stroke, as well as for aICAS and aECAS^7^. In a hospital-based multi-center case-control study, Li et al. find that with every increase of 10 mmHg in office systolic BP (SBP) and diastolic BP (DBP), the odds of ICAS increase by 32% and 28%^8^. In our previous study, we report a significant association of ambulatory early morning SBP with aICAS in patients with hypertension, with a 10-mmHg increment of SBP, the risk of aICAS increases by 30%^9^. Both ambulatory and HBP monitoring (HBPM), are recognized as important and practical tools to detect and manage hypertension, and to monitor BP variability (BPV)^10^, among which HBPM could provide BP values that are more relevant to daily-life circumstances. However, although amounts of works have illustrated the predictive value of HBP and HBPV for cardiovascular events and mortality^11^, few studies focus on the relationships between HBP and aICAS or aECAS. A recent study performed in general Japanese men finds the association of aICAS with HBP value but not HBPV^12^. Therefore, in the present study, we perform an association study in Chinese hypertension patients, aiming to analyze the effect of HBP and HBPV on both aICAS and aECAS, which were assessed by computed tomography angiography (CTA).

## Methods

### Study population

This study was undertaken within the framework of an ongoing cross-section and prospective study in China, which was a CTA-based study of aICAS and aECAS in stroke-free patients, who suffered from hypertension. The details and methodology have been previously reported^13^. Between May 2017 and November 2020, we performed the follow-up study on subjects recruited between May 2012 and December 2015, which were outpatients in Xinzhuang Community Hospital and then scheduled for a visit to Ruijin Hospital, a general hospital in Shanghai. The subjects at baseline were included if they were willing to take brain CTA, guaranteed long-term follow-up, and had at least two cardiovascular risk factors as defined in Chinese hypertension guidelines^14^. Subjects were excluded if they had a previous stroke, transient ischemic attack, atrial fibrillation, or iodine allergy.

### Ethics statement

The study was performed according to the guidelines of the Helsinki Declaration and was approved by the Ethics Committee of Ruijin Hospital. Written informed consent was obtained from all participants.

### HBP measurements

HBP was self-measured using a validated, memory-equipped, automatic oscillometric device (Omron HEM-7080IC; Omron Corp, Tokyo, Japan), which was able to transfer the readings to the computer via a universal serial USB connection. Patients received oral and written instructions on how to conduct HBP monitoring correctly. They were instructed to take three measurements with a 1-minute interval, each morning (4:00-10:00 am, just before taking antihypertensive drugs and breakfast, after urination) and evening (6:00-12:00 pm, before sleep) for a period of 7 consecutive days, whereas relaxed in the sitting position with back and arm supported and legs uncrossed for 5 minutes. All the HBP readings with date and hour were downloaded after the patients returned the device. Among the 627 patients enrolled in the study, 534 patients who completed HBP measurements on at least 3 days and measured at least 2 consecutive measurements every morning and evening^15^, were included in the final analysis. For separate and conjunct analysis of morning and evening self-measured BP at home, the average of corresponding readings was considered as morning or evening HBP. HBPV was quantified by calculating the standard deviation (SD), coefficient of variation (CV), and maximum-minimum difference (MMD) of morning or evening HBP between the mean BP across 7 days, respectively. Only systolic blood pressure (SBP) was included in the analysis as it is a stronger risk factor for stroke than diastolic blood pressure (DBP)^16^.

### CTA examination

A 64-section helical CT scanner (GE FX/I, General Electric, Fairfield, CT) as previously described^17^, was used to perform brain CTA. CTA acquisitions of every patient were obtained after a single bolus intravenous injection of 70 ml Optiray Ioversol 320 into the antecubital vein at a rate of 3 ml/sec. Scanning covered the whole brain down to the level of the aortic arch with a 5-mm slice thickness. Images were reformatted in axial, sagittal, and coronal planes with 1.25-mm slice thickness. All images were read by two experienced radiologists who were blinded to the clinical data of the patients. Stenosis was defined as a lesion that decreased arterial internal diameter and the worst stenosis at a cerebral artery was chosen as the representative for each subject. The percentage of stenosis was calculated as the ratio of the diameter of the diseased artery at its most severe site divided by the diameter of a nearby normal segment. The number of arteries with stenosis for each patient was also counted. The intracranial arteries included the intracranial segment of the internal carotid artery and vertebral artery, basilar artery, anterior cerebral artery, middle cerebral artery, and posterior cerebral artery. The extracranial arteries included the extracranial segment of the internal carotid artery and vertebral artery, external carotid artery, common carotid artery, and subclavian artery. The two radiologists had a good agreement in the designation of stenosis (κ = 0.93, P < 0.001). All disagreements were reviewed and adjudicated by a senior radiologist to reach a consensus. According to the presence of intra- and extracranial stenosis, patients were divided into four groups: no stenosis, isolated ICAS, isolated ECAS, and concurrent intra- and extracranial stenosis.

### Demographic and clinical measurements

Clinical information about the history of diabetes, current drug intake, smoking and drinking, and so on was collected by questionnaires in accordance with the standard procedure. Current smokers were defined as those who had smoked cigarettes on one or more days in the past 30 days. Body mass index (BMI) was body weight in kilograms divided by height in meters squared, and body weight and height were both recorded with participants wearing light indoor clothing and no shoes. Low-density lipoprotein cholesterol (LDL), fasting blood glucose, and serum creatinine were performed in the Central Laboratory of Ruijin Hospital (Shanghai, China) using the standard protocols.

### Statistical analysis

For database management and statistical analysis, we used R software (version 4.0.2, R core team, Vienna, Austria) with the R package base, nnet, and lmtest. Quantitative descriptive statistics were compared between patients in accordance with the presence of aICAS/aECAS by using ANOVA, and categorical variables were compared using the chi-square test. Multivariate-adjusted multinomial logistic regression analyses were used to examine the association of aICAS/aECAS with HBP indexes. Participants without stenosis were taken as the reference group. Separate analyses were performed for morning and evening home SBP, and for different HBPV indexes, adjusting for age, sex, BMI, LDL, smoking history, alcohol consumption, history of diabetes, and antihypertensive treatment. In further analyses, home SBP was forced into the model including corresponding HBPV. Improvement in the fit of logistic regression models was assessed using the log-likelihood ratio. Statistical significance was a 2-tailed α of ≤ 0.05.

## Results

### Participants characteristics

The clinical characteristics of the study participants were presented according to the presence or absence of aICAS/aECAS in Table 1. Among 534 patients, 116 had isolated aICAS, 73 had isolated aECAS, and 168 had concurrent aICAS and aECAS. Subjects without intra- or extracranial stenosis were significantly younger and with lower fasting blood glucose, while subjects with concurrent ICAS and ECAS were more likely to suffer a history of diabetes. Among 534 participants, a relatively high proportion (471, 88.2%) had received antihypertensive treatment. Moreover, subjects with concurrent aICAS and aECAS were with higher HBP and HBPV than the other three groups.

**Table 1.**
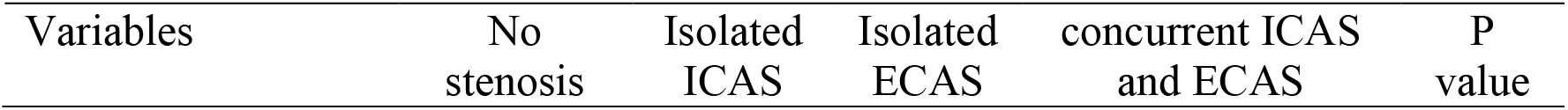
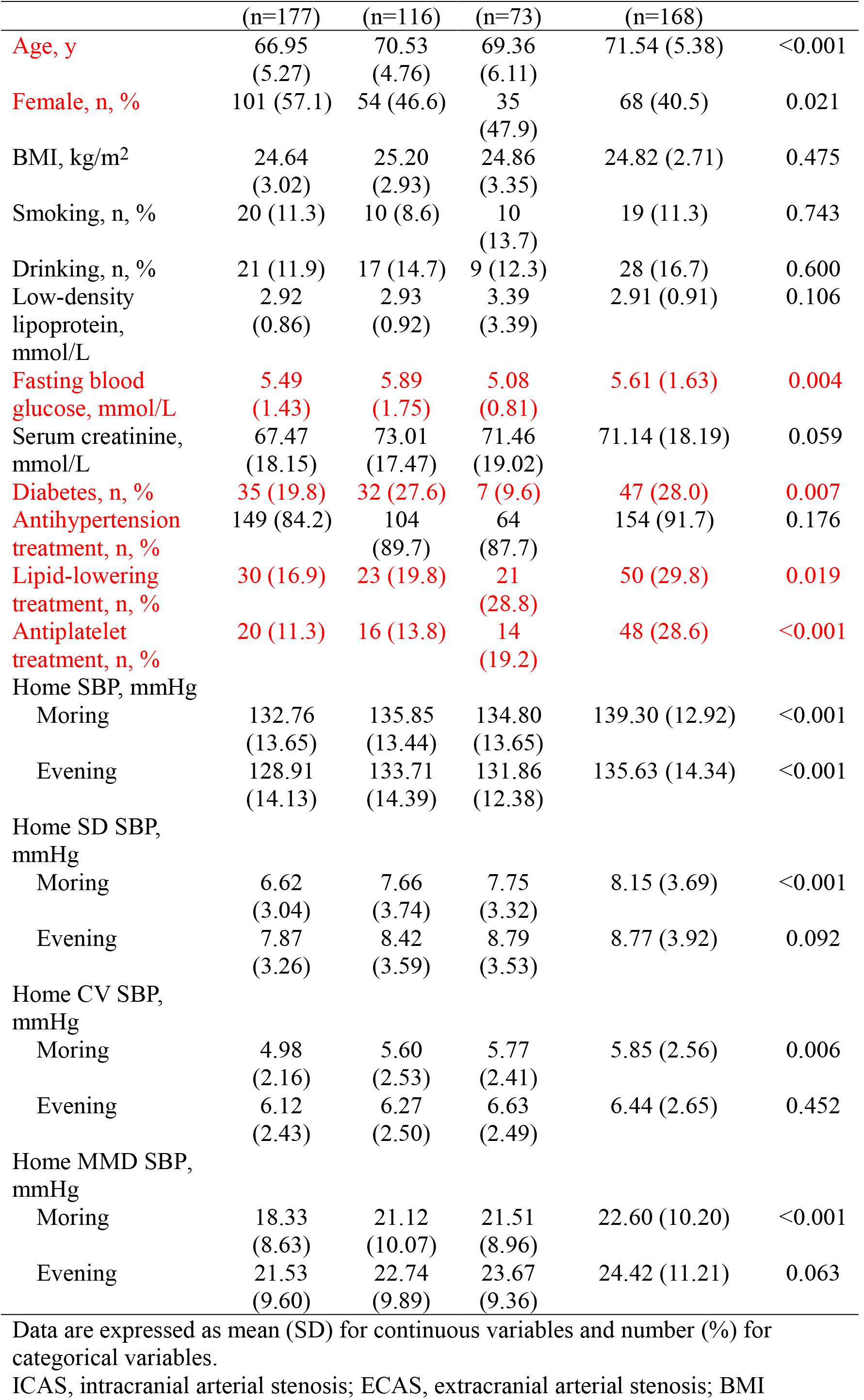
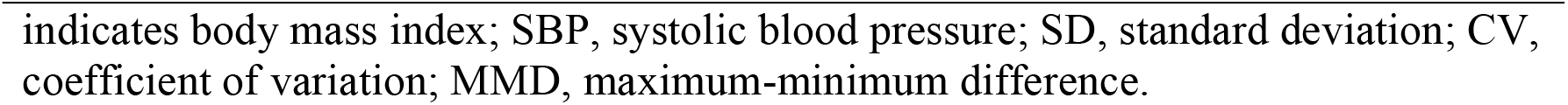
The characteristics of participants according to with or without intracranial and extracranial arterial stenosis.

| Variables | No stenosis | Isolated ICAS | Isolated ECAS | concurrent ICAS and ECAS | P value |
| --- | --- | --- | --- | --- | --- |

|  | (n=177) | (n=116) | (n=73) | (n=168) |  |
| --- | --- | --- | --- | --- | --- |
| Age, y | 66.95<br>(5.27) | 70.53<br>(4.76) | 69.36<br>(6.11) | 71.54 (5.38) | <0.001 |
| Female, n, % | 101 (57.1) | 54 (46.6) | 35<br>(47.9) | 68 (40.5) | 0.021 |
| BMI, kg/m<sup>2</sup> | 24.64<br>(3.02) | 25.20<br>(2.93) | 24.86<br>(3.35) | 24.82 (2.71) | 0.475 |
| Smoking, n, % | 20 (11.3) | 10 (8.6) | 10<br>(13.7) | 19 (11.3) | 0.743 |
| Drinking, n, % | 21 (11.9) | 17 (14.7) | 9 (12.3) | 28 (16.7) | 0.600 |
| Low-density lipoprotein, mmol/L | 2.92<br>(0.86) | 2.93<br>(0.92) | 3.39<br>(3.39) | 2.91 (0.91) | 0.106 |
| Fasting blood glucose, mmol/L | 5.49<br>(1.43) | 5.89<br>(1.75) | 5.08<br>(0.81) | 5.61 (1.63) | 0.004 |
| Serum creatinine, mmol/L | 67.47<br>(18.15) | 73.01<br>(17.47) | 71.46<br>(19.02) | 71.14 (18.19) | 0.059 |
| Diabetes, n, % | 35 (19.8) | 32 (27.6) | 7 (9.6) | 47 (28.0) | 0.007 |
| Antihypertension treatment, n, % | 149 (84.2) | 104<br>(89.7) | 64<br>(87.7) | 154 (91.7) | 0.176 |
| Lipid-lowering treatment, n, % | 30 (16.9) | 23 (19.8) | 21<br>(28.8) | 50 (29.8) | 0.019 |
| Antiplatelet treatment, n, % | 20 (11.3) | 16 (13.8) | 14<br>(19.2) | 48 (28.6) | <0.001 |
| Home SBP, mmHg |  |  |  |  |  |
| Morning | 132.76<br>(13.65) | 135.85<br>(13.44) | 134.80<br>(13.65) | 139.30 (12.92) | <0.001 |
| Evening | 128.91<br>(14.13) | 133.71<br>(14.39) | 131.86<br>(12.38) | 135.63 (14.34) | <0.001 |
| Home SD SBP, mmHg |  |  |  |  |  |
| Morning | 6.62<br>(3.04) | 7.66<br>(3.74) | 7.75<br>(3.32) | 8.15 (3.69) | <0.001 |
| Evening | 7.87<br>(3.26) | 8.42<br>(3.59) | 8.79<br>(3.53) | 8.77 (3.92) | 0.092 |
| Home CV SBP, mmHg |  |  |  |  |  |
| Morning | 4.98<br>(2.16) | 5.60<br>(2.53) | 5.77<br>(2.41) | 5.85 (2.56) | 0.006 |
| Evening | 6.12<br>(2.43) | 6.27<br>(2.50) | 6.63<br>(2.49) | 6.44 (2.65) | 0.452 |
| Home MMD SBP, mmHg |  |  |  |  |  |
| Morning | 18.33<br>(8.63) | 21.12<br>(10.07) | 21.51<br>(8.96) | 22.60 (10.20) | <0.001 |
| Evening | 21.53<br>(9.60) | 22.74<br>(9.89) | 23.67<br>(9.36) | 24.42 (11.21) | 0.063 |

Data are expressed as mean (SD) for continuous variables and number (%) for categorical variables.
ICAS, intracranial arterial stenosis; ECAS, extracranial arterial stenosis; BMI
indicates body mass index; SBP, systolic blood pressure; SD, standard deviation; CV, coefficient of variation; MMD, maximum-minimum difference.

### Association of aICAS and aECAS with HBP indexes

Table 2 exhibited the association of HBP and HBPV with aICAS/aECAS estimated by multivariate-adjusted multinomial logistic regression. The increased morning HBPV was significantly associated with the presence of isolated aICAS and aECAS after adjusting for age, sex, BMI, LDL, smoking history, alcohol consumption, history of diabetes, and antihypertensive treatment. Each 1-mmHg increment of HBPV increased by 10% (95% CI, 1.01-1.19; P = 0.021) odds for isolated aICAS, 14% (95% CI, 1.04-1.25; P=0.004) for isolated aECAS, and 14% (95% CI, 1.06-1.23; P<0.001) for concurrent aICAS and aECAS, respectively. Similar results were found in the analyses according to CV and MMD of morning HBP. However, neither morning nor evening HBP showed association with isolated aICAS or aECAS. Even additional adjustment for morning HSBP, the correlation of morning HBP SD with isolated ICAS (OR, 1.10; 95% CI, 1.01-1.19; P=0.025), isolated aECAS (OR, 1.14; 95% CI, 1.04-1.25; P=0.004) remained statistically significant.

**Table 2.**
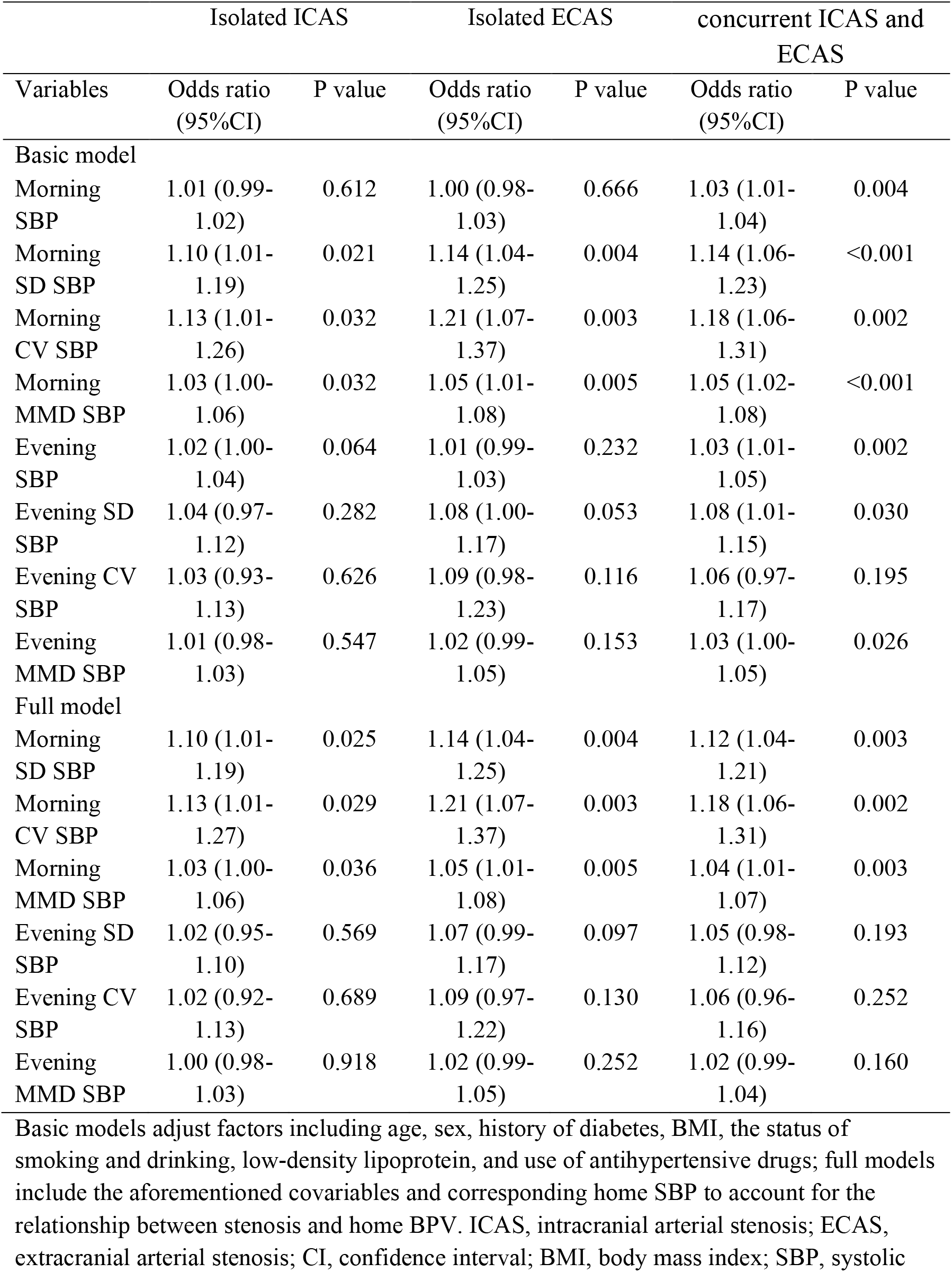
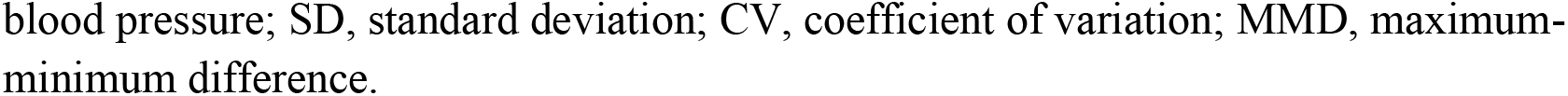
Associations of home SBP and variability with asymptomatic intra- and extracranial arterial stenosis.

| Variables | Isolated ICAS |  | Isolated ECAS |  | concurrent ICAS and ECAS |  |
| --- | --- | --- | --- | --- | --- | --- |
|  | Odds ratio (95%CI) | P value | Odds ratio (95%CI) | P value | Odds ratio (95%CI) | P value |
| Basic model |  |  |  |  |  |  |
| Morning SBP | 1.01 (0.99-1.02) | 0.612 | 1.00 (0.98-1.03) | 0.666 | 1.03 (1.01-1.04) | 0.004 |
| Morning SD SBP | 1.10 (1.01-1.19) | 0.021 | 1.14 (1.04-1.25) | 0.004 | 1.14 (1.06-1.23) | <0.001 |
| Morning CV SBP | 1.13 (1.01-1.26) | 0.032 | 1.21 (1.07-1.37) | 0.003 | 1.18 (1.06-1.31) | 0.002 |
| Morning MMD SBP | 1.03 (1.00-1.06) | 0.032 | 1.05 (1.01-1.08) | 0.005 | 1.05 (1.02-1.08) | <0.001 |
| Evening SBP | 1.02 (1.00-1.04) | 0.064 | 1.01 (0.99-1.03) | 0.232 | 1.03 (1.01-1.05) | 0.002 |
| Evening SD SBP | 1.04 (0.97-1.12) | 0.282 | 1.08 (1.00-1.17) | 0.053 | 1.08 (1.01-1.15) | 0.030 |
| Evening CV SBP | 1.03 (0.93-1.13) | 0.626 | 1.09 (0.98-1.23) | 0.116 | 1.06 (0.97-1.17) | 0.195 |
| Evening MMD SBP | 1.01 (0.98-1.03) | 0.547 | 1.02 (0.99-1.05) | 0.153 | 1.03 (1.00-1.05) | 0.026 |
| Full model |  |  |  |  |  |  |
| Morning SD SBP | 1.10 (1.01-1.19) | 0.025 | 1.14 (1.04-1.25) | 0.004 | 1.12 (1.04-1.21) | 0.003 |
| Morning CV SBP | 1.13 (1.01-1.27) | 0.029 | 1.21 (1.07-1.37) | 0.003 | 1.18 (1.06-1.31) | 0.002 |
| Morning MMD SBP | 1.03 (1.00-1.06) | 0.036 | 1.05 (1.01-1.08) | 0.005 | 1.04 (1.01-1.07) | 0.003 |
| Evening SD SBP | 1.02 (0.95-1.10) | 0.569 | 1.07 (0.99-1.17) | 0.097 | 1.05 (0.98-1.12) | 0.193 |
| Evening CV SBP | 1.02 (0.92-1.13) | 0.689 | 1.09 (0.97-1.22) | 0.130 | 1.06 (0.96-1.16) | 0.252 |
| Evening MMD SBP | 1.00 (0.98-1.03) | 0.918 | 1.02 (0.99-1.05) | 0.252 | 1.02 (0.99-1.04) | 0.160 |

Basic models adjust factors including age, sex, history of diabetes, BMI, the status of smoking and drinking, low-density lipoprotein, and use of antihypertensive drugs; full models include the aforementioned covariables and corresponding home SBP to account for the relationship between stenosis and home BPV. ICAS, intracranial arterial stenosis; ECAS, extracranial arterial stenosis; CI, confidence interval; BMI, body mass index; SBP, systolic
blood pressure; SD, standard deviation; CV, coefficient of variation; MMD, maximum-minimum difference.

Both morning and evening HBP and HBPV were independently related to concurrent aICAS and aECAS in separate regression analyses (P<0.05), except for CV with evening HBP. While compared to those without aICAS or aECAS, higher morning (95% CI, 1.00-1.04; P=0.035) and evening (95% CI, 1.01-1.04; P=0.008) HBP accounted for 2% odds increment of concurrent aICAS and aECAS. The identical outcomes were repeated for CV and MMD of morning HBP. With respect to evening HBPV, the relationship with concurrent ICAS and ECAS turned out to be clearly insignificant after additionally adjusting for evening HBP (P>0.05), whichever HBPV indexes were calculated.

The log-likelihood ratios (Table 3) confirmed that adding morning HBPV indexes to models including all confounding factors increased model fitness with P-values ranging from 0.007 to 0.002.

**Table 3.** Fit of multinomial logistic models of home SBP and variability with asymptomatic intra- and extracranial arterial stenosis.

| Models | -2log L | $\chi^2$ | P value |
| --- | --- | --- | --- |
| Basic model | 1323.86 |  |  |
| + Morning SBP | 1314.2 | 9.7 | 0.021 |
| + Morning SD SBP | 1308.8 | 15.1 | 0.002 |
| + Morning CV SBP | 1310.7 | 13.1 | 0.004 |
| + Morning MMD SBP | 1308.8 | 15 | 0.002 |
| + Evening SBP | 1313.5 | 10.3 | 0.016 |
| + Evening SD SBP | 1317.8 | 6.11 | 0.107 |
| + Evening CV SBP | 1320.7 | 3.15 | 0.369 |
| + Evening MMD SBP | 1318 | 5.89 | 0.117 |
| Basic model + Morning SBP | 1314.2 |  |  |
| + Morning SD SBP | 1302.1 | 12.1 | 0.007 |
| + Morning CV SBP | 1301.2 | 13 | 0.005 |
| + Morning MMD SBP | 1302.1 | 12.1 | 0.007 |
| Basic model + Evening SBP | 1313.5 |  |  |
| + Evening SD SBP | 1310.2 | 3.32 | 0.345 |
| + Evening CV SBP | 1310.7 | 2.8 | 0.423 |
| + Evening MMD SBP | 1310.4 | 3.11 | 0.374 |

Basic models adjust factors including age, sex, history of diabetes, BMI, the status of smoking and drinking, low-density lipoprotein, and use of antihypertensive drugs; BMI, body mass index; SBP, systolic blood pressure; SD, standard deviation; CV, coefficient of variation; MMD, maximum-minimum difference.

## Discussion

In the present study, we for the first time estimated the association of aICAS and aECAS with HBP and HBPV in stroke-free patients with hypertension. Morning HBPV was associated with isolated and concurrent aICAS and aECAS after adjusting for covariates, even after adjusting for the level of morning HBP.

In accordance with our results, a piece of recent research in the general population shed light on that each 17.7 mmHg increase in morning HBP corresponded to an odds ratio of 1.33 for subclinical intracranial cerebrovascular disease^18^. As for asymptomatic extracranial cerebrovascular disease, a nationwide study from Japan has demonstrated an increment of morning HBP was related to extracranial cerebrovascular damage^19^. However, a significant association of evening home SBP with stenotic lesions, to some extent, was not detected in this study. Although the overwhelming viewpoint is that subjects with higher morning home SBP, instead of higher evening home SBP, are more likely to suffer from cardiovascular diseases^20, 21^, our observation suggested that both morning and evening HBP had an influence on intra- and extracranial arteries of subjects with hypertension. Actually, research focused on the relationship of evening home SBP with subclinical cerebrovascular disease was lacking, and more evidence is needed.

It was recognized that higher BPV from visit-to-visit or within 24-h, gave rise to a more severe cerebrovascular damge^22^. When it comes to HBPV, previous studies have mainly concentrated on the adverse effects of increased HBPV on the stiffness of the large arteries, aggravating cardiovascular disease burden^23, 24^. In agreement with the present studies, some existing research has revealed the positive correlation between HBPV and isolated aECAS^25, 26^. Unfortunately, the clinical relevance of mid-term BPV in asymptomatic individuals with ICAS remains controversial. A recent study involving 677 healthy men determined that there was no significant association between HBPV and aICAS^12^. On the contrary, the key finding to emerge from our study was that morning HBPV was confirmed to increase the odds of isolated aICAS by 3 to 13 percent in stroke-free patients with hypertension. A possible explanation for the disparity might owe to different characteristics of the study populations. For example, a larger proportion of our participants have received anti-hypertension treatment, which have a higher morning HBPV in our research. Incidentally, we speculated that HBPV may reflect the increased stiffness and decreased compliance of the large elastic arteries, leading to endothelial dysfunction and inflammation, which accompany the growth of cerebral atherosclerosis^25, 27, 28^. Considering stenoses developing and progressing silently over the years, thus, the importance of paying close attention to HBPV of hypertensive patients was worth being stressed.

Limitations are acknowledged in this study and go as follows. Firstly, further validations are supposed to be undertaken in prospective cohort studies or randomized controlled trials for it is a cross-sectional study. Secondly, considering the disparity in the prevalence of aICAS and aECAS in different ethnic populations, discrepant results may occur. Thirdly, with a relatively small sample size, findings from this study should be interpreted with caution. Finally, information on participants’ sleeping patterns was not collected, as well as office BP.

In conclusion, the findings of this study confirmed the strong correlation of aICAS/aECAS with HBP in the hypertensive population, particularly morning HBPV. In the era of personalized medicine, these relations are well worth further investigation to mitigate the staggering burden of stroke posed at both individual and societal levels, with the widespread application of out-of-office measurements, especially cost-effective monitoring at home.

## Data Availability

No additional data are available.

## Contributors

EC and DA, JZ, XT, and YL are joint first authors. DZ and YW obtained funding. YL, DZ, and YW designed the study. EC, DA, JZ, XT, YW, and YL collected the data. EC and WY were involved in data cleaning, mortality follow-up, and verification. EC and DA analyzed the data. EC drafted the manuscript. YW and DZ contributed to the interpretation of the results and critical revision of the manuscript for important intellectual content and approved the final version of the manuscript. All authors have read and approved the final manuscript.

